# Indole 3-acetate and response to therapy in borderline resectable or locally advanced pancreatic cancer

**DOI:** 10.1101/2024.01.03.24300751

**Authors:** Peder R. Braadland, Ingvild Farnes, Elin H. Kure, Sheraz Yaqub, Adrian McCann, Per M Ueland, Knut Jørgen Labori, Johannes R. Hov

**Affiliations:** Research Institute of Internal Medicine and Norwegian PSC Research Center, Division of Surgery, Inflammatory Diseases and Transplantation, Oslo University Hospital, Oslo, Norway; Institute of Clinical Medicine, University of Oslo, Oslo, Norway; Department of Hepato-Pancreato-Biliary Surgery, Oslo University Hospital, Rikshospitalet, Oslo, Norway; Department of Cancer Genetics, Institute for Cancer Research, Oslo University Hospital, Oslo, Norway; Department of Natural Sciences and Environmental Health, University of South-Eastern Norway, Bø i Telemark, Norway; BEVITAL AS, Bergen, Norway; Department of Clinical Science, University of Bergen, Bergen, Norway; Section of Gastroenterology, Department of Transplantation Medicine, Oslo University Hospital, Oslo, Norway

## Abstract

**Background:** Recently, Tintelnot *et al* reported that a higher concentration of the bacterially produced metabolite indole 3-acetate (3-IAA) in blood was linked to a better response to chemotherapy in people with metastatic pancreatic ductal adenocarcinoma (PDAC). Here, we aimed to extend these observations to people diagnosed with non-metastatic PDAC.

**Patients and methods:** We measured circulating 3-IAA in samples from a prospective population-based cohort of 124 individuals with borderline resectable or locally advanced PDAC who later underwent primary chemotherapy. The majority (61%) of the individuals were treated with FOLFIRINOX. We used univariable and multivariable Cox proportional hazards regression to estimate the association between pre-treatment 3-IAA and overall survival.

**Results:** The median serum 3-IAA concentration before chemotherapy was 290 (interquartile range 203–417) ng/mL. The unadjusted hazard ratio (HR) for pre-treatment log_2_(3-IAA) was 0.93, 95% confidence interval (CI) [0.74–1.16], p=0.52. When adjusting for age, ECOG, CA19-9 and tumor classification, the HR for log_2_(3-IAA) was 0.87, 95% CI [0.68–1.12], p=0.28.

**Conclusions:** Our findings suggest that the potentiating effect of 3-IAA observed in metastatic PDAC may not translate to borderline resectable or locally advanced PDAC. We recommend additional clinical validation of 3-IAA’s predictive value before implementation attempts in human studies are initiated.

## Introduction

Biomarkers of chemotherapy response are needed to increase response rates and personalize treatment. Recently, Tintelnot et al.^1^ identified circulating indole 3-acetate (3-IAA) as positively associated with response to chemotherapy in metastatic pancreatic ductal adenocarcinoma (PDAC). The molecular mechanism depended on oxidation of 3-IAA by myeloperoxidase from neutrophils and subsequent increased reactive oxygen species in tumor cells. The study introduced a new concept of a microbial metabolite modifying chemotherapy response,^2^ which could even be improved by supplementing the diet with the 3-IAA precursor tryptophan in mice.

In borderline resectable or locally advanced PDAC, systemic chemotherapy is the primary initial treatment strategy to facilitate resection with curative intent.^3^ We therefore measured 3-IAA in a cohort of people with borderline resectable or locally advanced PDAC before starting chemotherapy,^4^ aiming to validate the observations by Tintelnot *et al*. in a PDAC population without distant metastases.

## Patients and methods

### Study design and participants

The participants included in this study were from the NORPACT2 study,^4^ which was a prospective, population-based cohort of people with borderline resectable or locally advanced PDAC enrolled at the Oslo University Hospital between 2018 and 2021. We included 124 individuals, who donated blood before initiation of any chemotherapy. A subset of 35 (28%) had a second blood draw 2–4 months after initiation of chemotherapy.

### Targeted liquid chromatography-tandem mass spectrometry (LC-MS/MS)

Sample handling and the LC-MS/MS procedure is detailed in the online-only data.

### Outcome, definitions and diagnostic criteria

Borderline resectable or locally advanced PDAC was diagnosed according to the National Comprehensive Cancer Network (NCCN) criteria, version 2, 2017.^3^ Distant metastases were ruled out using computed tomography (CT) scans of the abdomen and chest. Fine-needle aspiration cytology or biopsy by endoscopic ultrasound was required to confirm PDAC. The chemotherapy regimen was decided by the treating medical oncologist at the local hospital.

The primary outcome was defined as overall survival, and time to the outcome was defined as the time from blood draw, which was done in conjunction with initiation of primary chemotherapy. A secondary outcome was included where participants were censored at their time of surgical resection (after start of primary chemotherapy) where applicable.

### Statistics

Surviving proportions were visualized by plotting Kaplan-Meier survival curves of quartiles of 3-IAA. Cox proportional hazards models were used to estimate 3-IAA’s (log_2_-transformed) association with overall survival. We included ECOG performance status (modeled as a continuous variable), CA19-9 (log_2_-transformed), tumor classification (locally advanced or borderline resectable)) and age in a multivariable Cox model.

To test for differences in distributions of continuous variables we used the Wilcoxon rank-sum tests for independent or paired observations. Distributions are given as median (interquartile range, IQR).

For any variable used in the analyses, data were missing in less than 5% of the participants and therefore not imputed. All data processing, visualization and statistical analyses were performed in R.

## Results

In total, 124 individuals (median 68 years, 47% male) with borderline resectable (n=68) or locally advanced PDAC (n=56) were included (**Table 1**). Seventy-six (61%) individuals were treated with fluorouracil, leucovorin, irinotecan, and oxaliplatin (FOLFIRINOX), 26 (21%) with gemcitabine plus nab-paclitaxel, and 22 (18%) with gemcitabine monotherapy or other chemotherapies.

**Table 1.**
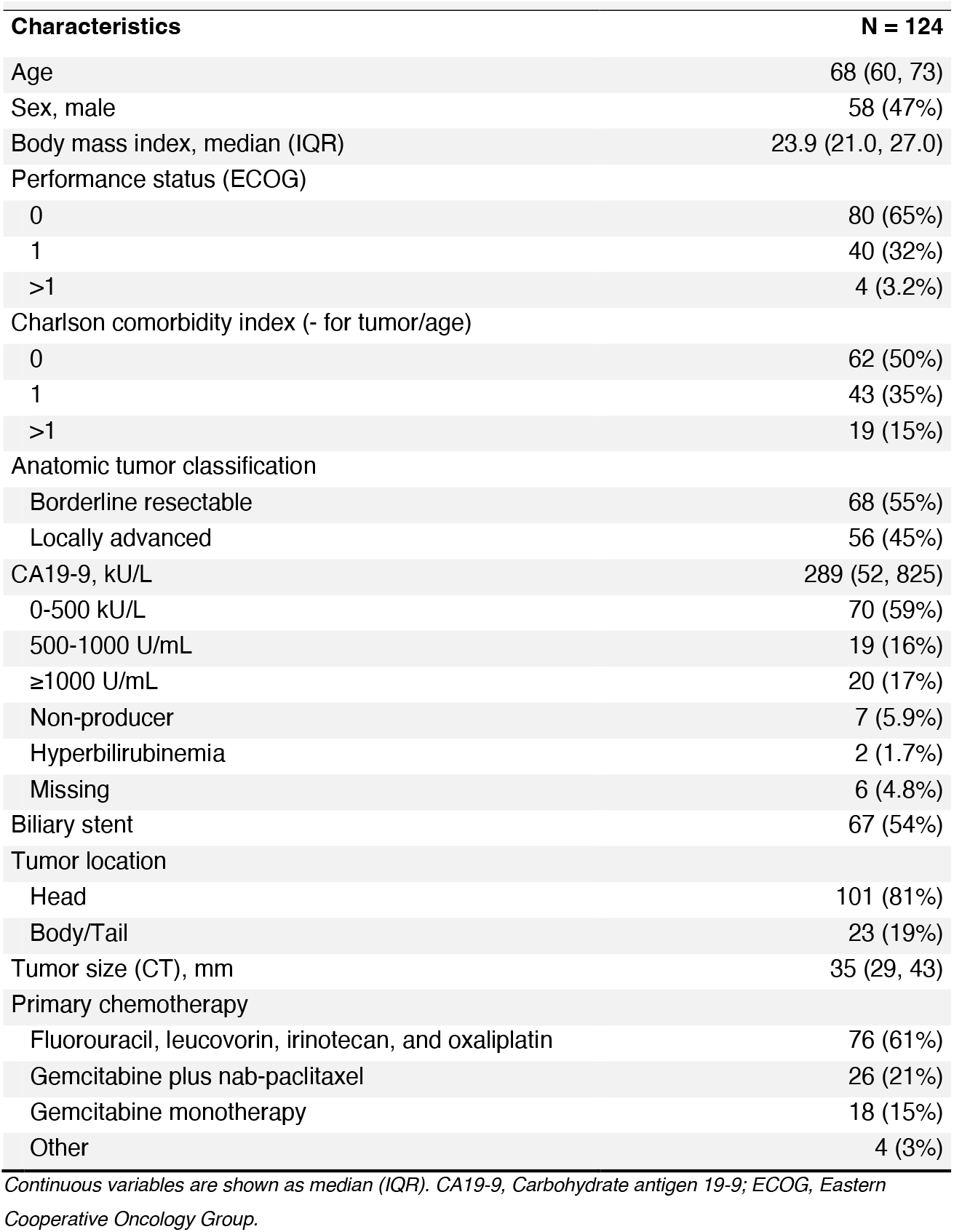
Baseline characteristics of the study population.

The median 3-IAA concentration before chemotherapy was 290 (IQR 203–417) ng/mL. Serum 3-IAA was positively correlated with age (Spearman’s ρ=0.33, p<0.001; Figure S1a) but not with tumor diameter or serum CA19-9 (Figure S1b and c). Pre-treatment 3-IAA was similar in the different chemotherapy groups (Figure S1d).

We found no statistically significant differences in pre-treatment 3-IAA concentrations according to response as defined by CT scan or CA19-9 decline (Figure 1a, b). 3-IAA concentration was numerically higher in individuals who later had surgical resection and in those alive after one year, but the observations were not statistically significant (Figure 1c, d). In participants with a second sample taken, 3-IAA concentrations were on average increased after chemotherapy (median 265 nmol/L before and 328 nmol/L after), but the increase was not statistically significant (p=0.24).

**Figure 1.**
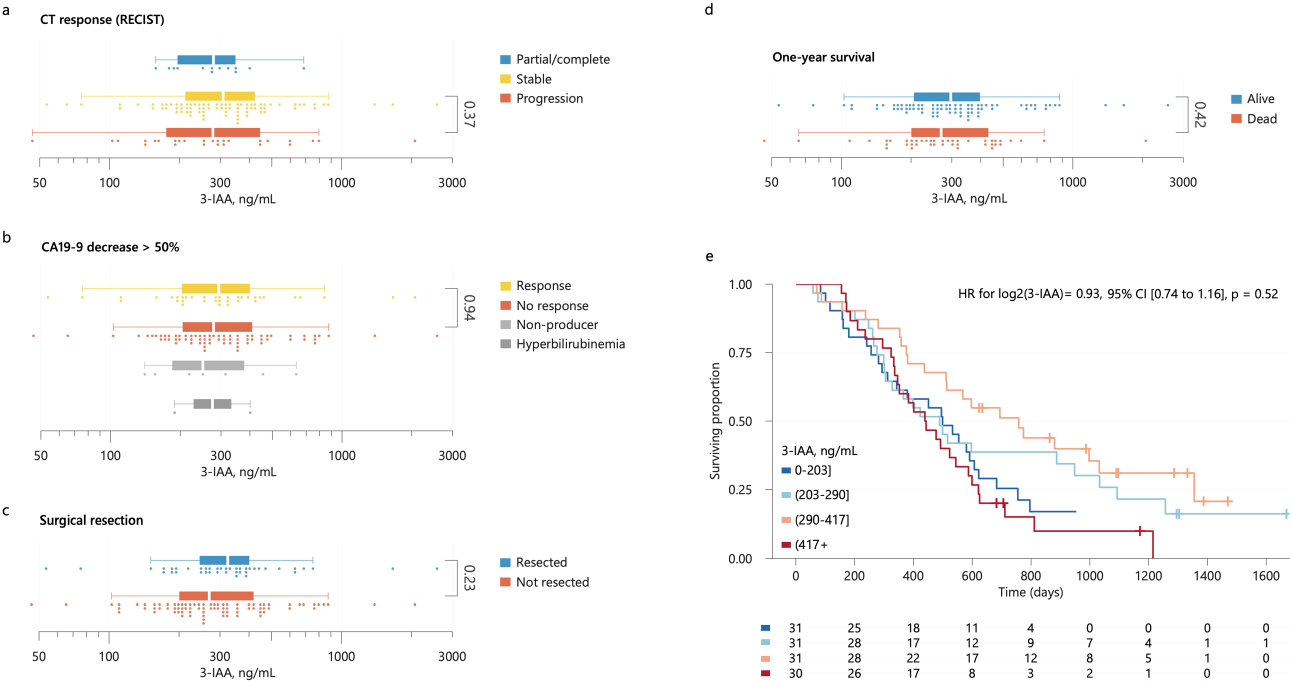
**a-d**. Distributions of 3-IAA by CT-response according to RECIST criteria (a), CA19-9 decrease > 50% (b), whether surgical resection after neoadjuvant chemotherapy was performed or not (c), and by one year survival status (d). Statistical significance was tested using Mann-Whitney U tests. **e**. Kaplan-Meier survival curves for overall survival for participants categorized by their pre-treatment 3-IAA concentration (quartiles) with number at risk below the plot. The result from a univariable Cox model for log(3-IAA) is shown as inset in the plot.

The median survival time was 499 days (95% confidence interval (CI) 437–597). Twenty-six (21%) individuals were event-free at their last follow-up date. We stratified participants into quartiles by pre-treatment 3-IAA concentrations but found no significant differences in survival between the groups (Figure 1e), a finding supported by a Cox model where 3-IAA (log_2_) had a HR=0.93, 95% CI [0.74–1.16], p=0.52. The model estimates were similar in the subgroups of borderline resectable or locally advanced cancer. Since 41 (33%) of the participants eventually had surgical resection of their primary tumor, which could influence survival, we evaluated the prognostic value of 3-IAA when censoring these cases at their time of surgery, with no evidence of differences in survival (Figure S2). Finally, using the full cohort, we adjusted 3-IAA for age, ECOG, CA19-9 and tumor classification, where 3-IAA (log_2_) had an adjusted HR=0.87, 95% CI [0.68–1.12], p=0.28.

## Discussion

In the present study, serum 3-IAA concentration was not associated with response to chemotherapy or overall survival in a cohort of borderline resectable or locally advanced PDAC starting chemotherapy. The results suggest that the previous findings of better chemotherapy response in people with metastatic PDAC and high 3-IAA are unlikely to extend to the current population.

There are several potential explanations for this disappointing finding. Whereas Tintelnot *et al*. studied metastatic PDAC, our population had no signs of distant metastasis.^4^ People with non-metastatic cancers often have a smaller total tumor load, better functional status, and differences in tumor biology compared with metastatic cancers, all of which may influence the effect of chemotherapy and its modifiers.

The median 3-IAA concentration in the present study was about 10-fold higher than what was found in Tintelnot *et al*. In a healthy control population analyzed in the context of other ongoing studies, the median 3-IAA was 377 (IQR 303–464) ng/mL. This was higher than in the present PDAC cohort but within the same order of magnitude. We speculate that the discrepancy in 3-IAA concentrations arose from assay differences. Nonetheless, if inter-individual variation in 3-IAA measurements was consistent with the two methods, we would expect to find an association with survival in our cohort if it truly existed. In our univariable model, 3-IAA had a wide confidence interval (0.74–1.16). While this makes it challenging to confidently assert the presence or absence of an effect, any potential clinical significance appears most likely to be small.

Apparent predictive performances in discovery cohorts are often optimistic and models commonly perform worse in new populations for a variety of reasons^5^. In the study from Tintelnot *et al*. the effect observed in the second human PDAC cohort was weaker than in the first cohort (explained variance in the correlation between 3-IAA and survival time was R^2^=0.51 in a cohort from Hamburg and R^2^=0.24 in a cohort from Munich).

In conclusion, our results do not align with the optimism generated by Tintelnot *et al’s* study but provide valuable insights. Proof of an effect in humans could probably be best obtained in a controlled clinical intervention study aimed at increasing circulating 3-IAA as an adjuvant to chemotherapy. Based on the current knowledge, such a trial should focus on metastatic PDAC. The first step should however be to validate the association between 3-IAA and chemotherapy response technically and clinically in geographically distinct cohorts of metastatic PDAC.

## Acknowledgements

The study and co-workers were funded by research grants from the Regional Health Authorities of South-Eastern Norway (no. 2018088, 2019029, 2023018) and the Norwegian Cancer Society (198039-2018), as well as by the strategic research area “Personalized microbiota therapy in clinical medicine” at Oslo University Hospital. J.R.H. is in part funded by a grant from the European Research Council (no. 802544).

## Author contributions

S.Y., K.J.L. and J.R.H. conceived the project. P.R.B., E.H.K., S.Y., K.J.L. and J.R.H. participated in experimental design, execution and data analyses. I.F. and K.J.L. collected the patient cohort and clinical data. A.M. and P.M.U. performed biochemical analyses. P.R.B., S.Y., K.J.L. and J.R.H. wrote the first draft. K.J.L. and J.R.H. jointly supervised the study and contributed equally. P.R.B., I.F., E.H.K., S.Y., A.M., P.M.U., K.J.L. and J.R.H. critically revised the manuscript and approved of the final version.

## Data availability

Data are not deposited in a public repository due to data privacy regulations in Norway and lack of participant consent. However, data are available on request, if the aim of the analysis is covered by the consent signed by the participants, following an amendment to the ethics approval and a data transfer agreement.

## Online-only material

### Supplementary methods

#### Targeted liquid chromatography-tandem mass spectrometry

Baseline and follow-up serum samples were stored according to a standardized procedure at Oslo University Hospital and were analyzed using a targeted liquid chromatography-tandem mass spectrometry (LC-MS/MS) platform measuring B-vitamins and human and bacterial tryptophan metabolites at BEVITAL (www.bevital.no) as described in Midttun *et al*. ^6^

Briefly, 3-IAA was added to an established assay^5^ using indole-2,4,5,6,7-d_5_-3-acetic acid (3-IAAd_5_), obtained from C/D/N Isotopes Inc (Quebec, Canada), as internal standard. The retention times were 4.93 min (3-IAA) and 4.91 min (3-IAAd_5_). The analytes were detected in positive-ion multiple reaction monitoring (MRM) mode, using the ion-pairs of 176.2/130.2 m/z and 181.1/134.1 m/z for 3-IAA and 3-IAAd_5_, respectively.

### Supplementary figures and legends

**Figure S1.**
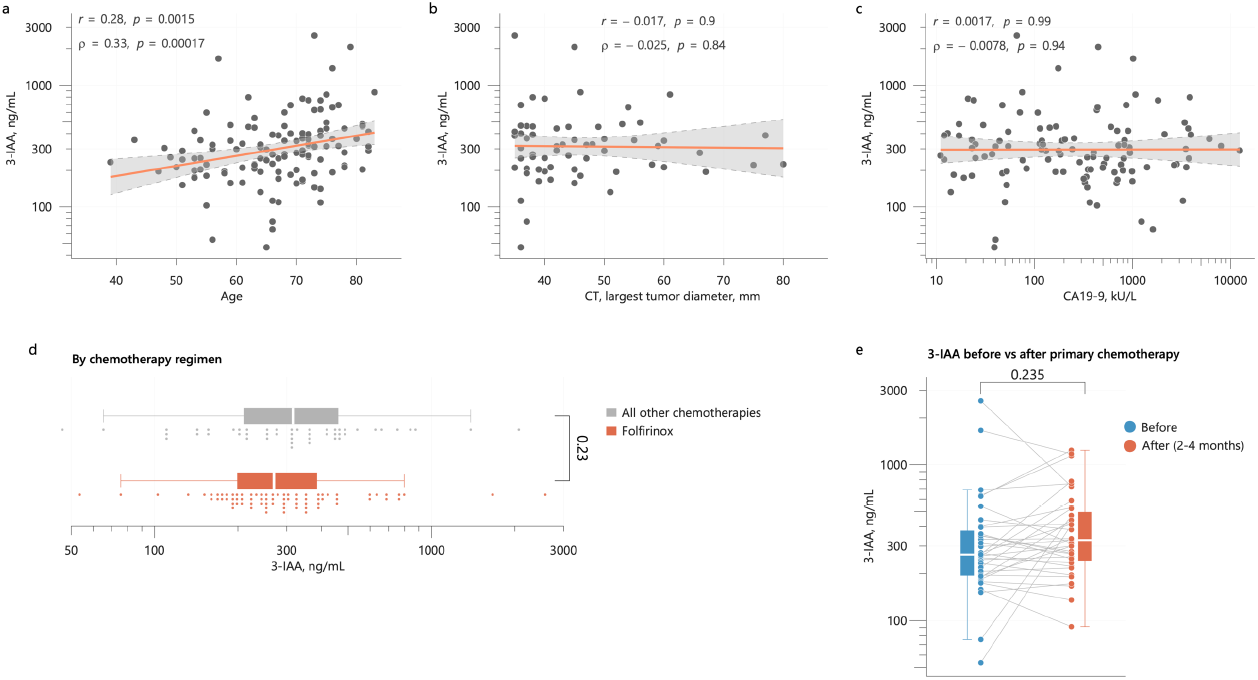
**a-c**. Plots of baseline 3-IAA concentrations versus age, largest tumor diameter (by CT-scan) and CA19-9, all measured at baseline (before primary chemotherapy). The bivariate associations were tested using Pearson’s (r) and Spearman’s (ρ) correlation. The red lines show the linear regression fits with shaded 95% confidence intervals. **d**. Distribution of pre-treatment 3-IAA by chemotherapy regimen (FOLFIRINOX versus all other). Statistical significance was tested using a Mann-Whitney U test. **e**. Distributions of 3-IAA before and after primary chemotherapy, with lines indicating changes from before to after. Statistical significance was tested using a paired test (Wilcoxon). 3-IAA = 3-indoleacetic acid, FFX = FOLFIRINOX.

**Figure S2.**
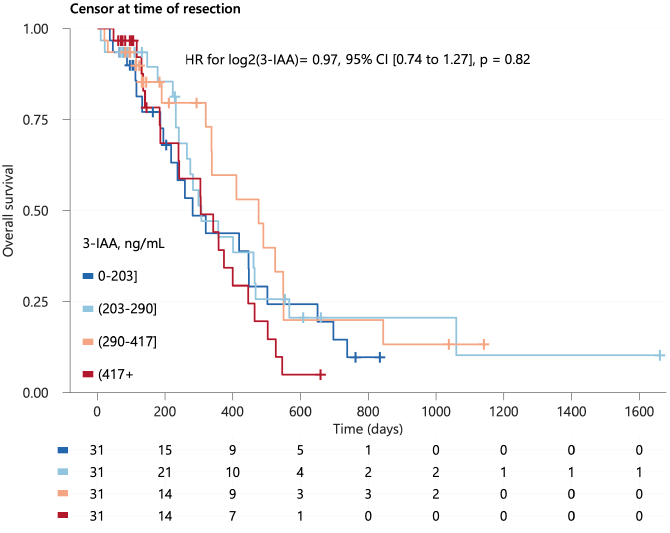
Kaplan-Meier survival curves for overall survival, censoring participants who underwent surgical resection at their times of surgery, categorized by their baseline 3-IAA concentration (quartiles) with number at risk below the plot. The results from a univariable Cox model for log2(3-IAA) is shown as an inset in the plot.

## References

1. Tintelnot J, Xu Y, Lesker TR, et al. Microbiota-derived 3-IAA influences chemotherapy efficacy in pancreatic cancer. Nature. 2023;615(7950):168–174. doi:10.1038/s41586-023-05728-y

2. Liu L, Shah K. The Potential of the Gut Microbiome to Reshape the Cancer Therapy Paradigm: A Review. JAMA Oncology. 2022;8(7):1059–1067. doi:10.1001/jamaoncol.2022.0494

3. Tempero MA, Malafa MP, Al-Hawary M, et al. Pancreatic Adenocarcinoma, Version 2.2021, NCCN Clinical Practice Guidelines in Oncology. J Natl Compr Canc Netw. 2021;19(4):439–457. doi:10.6004/jnccn.2021.0017

4. Farnes I, Kleive D, Verbeke C, et al. Resection rates and intention-to-treat outcomes in borderline and locally advanced pancreatic cancer - Real-world data from a population-based, prospective cohort study (NORPACT-2). BJS Open. 2023;7(6):zrad137. doi: 10.1093/bjsopen/zrad137.

5. Riley RD, Snell KI, Ensor J, et al. Minimum sample size for developing a multivariable prediction model: PART II - binary and time-to-event outcomes. Stat Med. 2019;38(7):1276–1296. doi:10.1002/sim.7992

6. Midttun Ø, Hustad S, Ueland PM. Quantitative profiling of biomarkers related to Bvitamin status, tryptophan metabolism and inflammation in human plasma by liquid chromatography/tandem mass spectrometry. Rapid Communications in Mass Spectrometry. 2009;23(9):1371–1379. doi:10.1002/rcm.4013

